# Determinants of mosaic chromosomal alteration fitness

**DOI:** 10.1101/2023.10.20.23297280

**Authors:** Yash Pershad, Taralynn Mack, Hannah Poisner, Yasminka A Jakubek, Adrienne M Stilp, Braxton D Mitchell, Joshua P Lewis, Eric Boerwinkle, Ruth J Loos, Nathalie Chami, Zhe Wang, Kathleen Barnes, Nathan Pankratz, Myriam Fornage, Susan Redline, Bruce M Psaty, Joshua C Bis, Ali Shojaie, Edwin K Silverman, Michael H Cho, Jeong Yun, Dawn DeMeo, Daniel Levy, Andrew Johnson, Rasika Mathias, Margaret Taub, Donna Arnett, Kari North, Laura M Raffield, April Carson, Margaret F Doyle, Stephen S. Rich, Jerome I. Rotter, Xiuqing Guo, Nancy Cox, Dan M Roden, Nora Franceschini, Pinkal Desai, Alex Reiner, Paul L Auer, Paul Scheet, Siddhartha Jaiswal, Joshua S Weinstock, Alexander G Bick

**Author notes:** Correspondence to: Dr. Alexander Bick, Vanderbilt University Medical Center, 550 Robinson Research Building, 2200 Pierce Ave, Nashville, TN 37232. Contributed equally (Yash Pershad, Taralynn Mack).

## Abstract

Clonal hematopoiesis (CH) is characterized by the acquisition of a somatic mutation in a hematopoietic stem cell that results in a clonal expansion. These driver mutations can be single nucleotide variants in cancer driver genes or larger structural rearrangements called mosaic chromosomal alterations (mCAs). The factors that influence the variations in mCA fitness and ultimately result in different clonal expansion rates are not well-understood. We used the Passenger-Approximated Clonal Expansion Rate (PACER) method to estimate clonal expansion rate for 6,381 individuals in the NHLBI TOPMed cohort with gain, loss, and copy-neutral loss of heterozygosity mCAs. Our estimates of mCA fitness were correlated (R^2^ = 0.49) with an alternative approach that estimated fitness of mCAs in the UK Biobank using a theoretical probability distribution. Individuals with lymphoid-associated mCAs had a significantly higher white blood cell count and faster clonal expansion rate. In a cross-sectional analysis, genome-wide association study of estimates of mCA expansion rate identified *TCL1A*, *NRIP1*, and *TERT* locus variants as modulators of mCA clonal expansion rate.

## Introduction

With age, hematopoietic stem cells (HSCs) accumulate somatic mutations.^1^ While most of these mutations have little effect on cell fitness and become simply “passengers”, some mutations, called “drivers”, increase the fitness of a HSC and lead to clonal expansion. The phenomenon of clonal hematopoiesis (CH) occurs when a clone of cells is detectable without causing blood count abnormalities or hematologic malignancy.^2^ CH is common and increases with age. Previous studies have shown that CH significantly increases an individual’s risk of all-cause mortality and many common complex diseases.^3–11^

Driver mutations in HSCs can be either single nucleotide variants (SNVs) in genes associated with hematological malignancies (e.g., *DNMT3A, TET2,* and *JAK2*)^12^ referred to as Clonal Hematopoiesis of Indeterminate Potential (CHIP) or larger chromosomal rearrangements called mosaic chromosomal alterations (mCAs).^9^ mCAs, which may involve a gain or loss of an entire chromosome or a copy-neutral loss of heterozygosity (CN-LOH), occur in about 3% of individuals older than 50 years old without cancer.^5,13^ Specific sets of mCAs have been associated with risk of lineage-specific hematologic malignancies.^14,15^ Although individuals with larger mCA clones generally have worse health outcomes, the factors that influence the variations in mCA fitness and ultimately result in different clonal expansion rates have not been studied and are not well-understood.^5,10,16,17^

Conventional methods to study clonal fitness require collecting serial blood samples over several decades, so obtaining sufficient sample sizes is challenging. To overcome this limitation, methods have been developed to estimate clonal fitness from a single blood draw. Watson and Blundell estimated fitness of SNVs and mCAs on a population level from clonal fraction distributions in UK Biobank participants.^18,19^ However, this method only predicts clonal expansion rate for a mutation aggregated over a population rather than for a mutation in an specific individual. Estimating the fitness of a given driver mutation for an individual is essential to elucidate germline modifiers of clonal expansion and better characterize the pathophysiology of CH-associated diseases.

We recently developed a method called passenger-approximated clonal expansion rate (PACER), which uses the abundance of passenger mutations accompanying a driver mutation to estimate the fitness of a SNV with a single blood sample from an individual.^20^ Here, we apply PACER to 6,381 individuals with gain, loss, and CN-LOH mCAs in the NHLBI Trans-Omics for Precision Medicine (TOPMed) dataset to identify determinants and consequences of mCA fitness (**Fig 1a**). PACER estimates of mCA fitness were compared to the fitness of SNV mutations implicated in CHIP. We examined associations between mCA expansion and peripheral blood counts and observed that for individuals with lymphoid-associated mCAs, faster expansion rates were associated with higher lymphocyte counts. Next, using our individual mCA fitness estimates, we performed a genome-wide association study (GWAS) and found that variants in *TCL1A*, *NRIP1*, and *TERT* may modulate mCA clonal expansion.

**Fig. 1:**
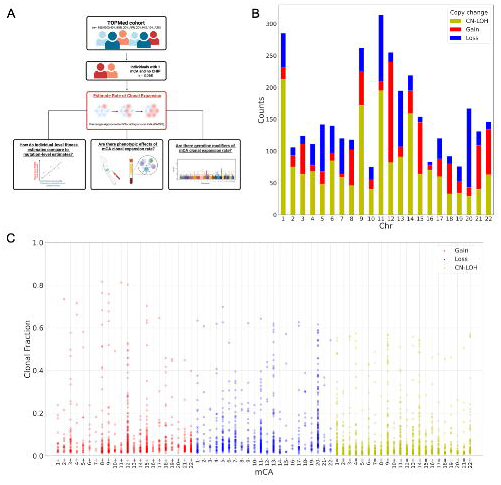
(**a**) Schematic of the study. Excluding mosaic chromosomal alterations (mCAs) of chromosome X, 3,068 individuals had 3,828 mCAs in TOPMed. 6,930 individuals had 1 mCA, and 763 had > 1 mCA. (**b**) Stacked bar plot showing counts of mCAs by chromosome, separated by copy change type: copy neutral loss of heterozygosity (CN-LOH) in yellow, gain in red, and loss in blue. mCAs of chromosome X were excluded. (**c**) Dot plot of clonal fractions for each patient with specific mCA (chromosome and copy change). Red and “+” represents gain of chromosome, blue and “-“ represents loss of chromosome, and yellow and “=” represents CN- LOH.

## Results

We identified 6,930 people with one mCA and 763 with multiple mCAs. After excluding mCAs of sex chromosomes, we detected 3,828 autosomal mCAs in 3,068 unique individuals (**Fig 1b**). The median age of individuals with autosomal mCAs was 67, and 57% were female. The most common autosomal mCAs were on chromosomes 11, 1, and 9, with CN-LOH being the majority (52%). Clonal fraction showed significant variation across autosomal mCA types and chromosomes (**Fig 1c**).

### Passenger-approximated clonal expansion rate of mCAs by chromosomal event

Leveraging the PACER method, we quantified mCA fitness in individuals with a single mCA and no CH-associated SNVs.^20^ Within this cohort, the minimum total passenger mutations for a patient with an mCA was 3, maximum was 933, and median was 53 (**Fig 2a**). We calculated a covariate-adjusted PACER for each individual from the normalized residuals of a negative binomial regression of age, sex, and clonal fraction predicting total passenger mutation count.

**Fig 2:**
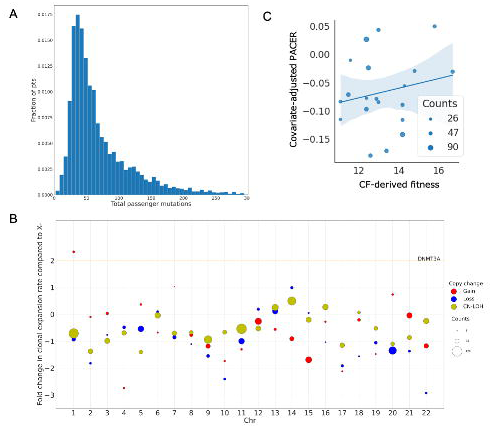
(**a**) Histogram of total passenger mutations for all individuals with 1 mosaic chromosomal abnormality in TOPMed. A passenger mutation is defined as a clocklike C>T or T>C substitution that does not occur in a CHIP-associated gene. (**b**) Dot plot of fold change in clonal expansion rate compared to loss of chromosome X (X-). Red dots represent a gain of chromosome, blue dots represent a loss of chromosome, and yellow dots represent CN-LOH. The clonal expansion rate is calculated after covariate adjustment (age, sex, study cohort, and clonal fraction) and inverse normalization of the total passenger mutations for a given individual. The fold change in clonal expansion rate is calculated by dividing the clonal expansion rate for a given mCA by that for a loss of chromosome X. The size of the dot corresponds to the number of individuals with that mCA type. The orange line represents the fold change of clonal expansion rate of the CHIP mutation DNMT3A with respect to X-. (**c**) Scatter plot of median total passenger mutations and fitness derived from clonal fraction by mCA (only includes mCAs with counts > 25). The size of the dot corresponds to the number of individuals with that mCA type. A generalized linear model of total passenger mutations, mean age, and mean clonal fraction predicting clonal-fraction-derived (CF-derived) fitness had an R^2^ value of 0.49.

Compared to SNVs, mCAs involve changes to larger regions of chromosomes. As a sensitivity analysis, we investigated if these larger rearrangements alter total passenger mutation counts, thereby confounding our estimations of fitness. To test this, we compared passenger mutation counts between mCA chromosomes and non-mCA chromosomes and found no significant difference by mCA location and type (**Extended Data Fig 1a**) or in aggregate (p = 0.12).

Furthermore, we found that PACER scores did not significantly change upon exclusion of the mCA chromosome. A linear regression of the covariate-adjusted PACER with and without the mCA chromosome demonstrated a high degree of correlation, with a Spearman’s rank correlation coefficient (ρ) of 0.82 (**Extended Data Fig 1b**). Moreover, a linear regression of covariate-adjusted PACER with and without a random chromosome had the same correlation, with a ρ of 0.82 (**Extended Data Fig 1c**).

We aggregated the individual data by mCA type and location and calculated the fold-change in estimated clonal expansion rate relative to a loss of chromosome X (**Fig 2b**). Gain of chromosome 1, gain of chromosome 7, loss of chromosome 14, and CN-LOH of chromosome 14 had the highest covariate-adjusted estimated clonal expansion rates. However, of those mCAs, only CN-LOH of chromosome 14 occurred in more than 25 individuals in TOPMed. Of all mCAs, only a gain in chromosome 1 had a higher covariate-adjusted estimated clonal expansion rate than driver SNVs in non-R882 *DNMT3A*.^20^

To validate our estimates, we compared our passenger-estimated fitness by mCA in TOPMed to those generated by another approach based upon clonal fraction distribution in the UK Biobank.^18^ A generalized linear model of median PACER and median age of individuals in TOPMed explained 49% of the variance in this clonal-fraction-derived (CF-derived) fitness in the UK Biobank (**Fig 2c**), compared to 14% for a model using only median age and 44% for a model using only median total passenger mutation counts. Median PACER had a significant positive association with CF-derived fitness (β = 0.04, 95% CI = [0.017, 0.063], p = 0.001).

We next investigated the fitness of two curated set of mCAs associated with future development of either myeloid or lymphoid hematologic malignancy.^14^ In TOPMed, the lymphoid set of mCAs had the highest covariate-adjusted PACER, followed by the myeloid mCAs and then mCAs associated with neither malignancy (**Extended Data Fig 2**). ANOVA demonstrated a significant difference between covariate-adjusted PACER among these three groups (p = 0.0048).

### mCA expansion rate associates with blood cell counts

To understand the relationship between clonal expansion rate with clinical phenotypes, we also investigated the association between clonal expansion rate and peripheral blood counts in individuals with mCAs. Blood counts were available for 987 individuals with mCAs. The median ± interquartile range (IQR) leukocyte count for individuals with mCAs was 6,400 ± 2,400 cells/µL. Using a curated set of mCAs associated with myeloid or lymphoid hematologic malignancy in other biobanks, individuals with lymphoid mCAs within TOPMed had a significantly (p = 0.0004) higher lymphocyte count (median ± IQR = 2,010 ± 1,060 cells/µL) than those with non-lymphoid mCAs (median ± IQR = 1,870 ± 850 cells/µL).^14,15^ Myeloid cell counts were not significantly different in individuals with myeloid mCAs compared to those with other mCAs (p = 0.544).

Among individuals with lymphoid mCAs, a multiple regression of age at time of blood draw, sex, clonal fraction, and PACER predicting lymphocyte count explained 13.3% of the variance in lymphocyte counts, compared to only 2.5% for a model without PACER (**Fig 3a**). In the multiple regression, PACER had a significant positive association with lymphocyte count (β = 0.0175, 95% CI [0.003, 0.032], p = 0.019). In contrast, for individuals with myeloid mCAs, the same regression explained 3.5% of the variance in myeloid cell counts; PACER was not associated with myeloid cell count (β = -0.0031, 95% CI [-0.009, 0.003], p = 0.298) (**Fig 3b**).

**Fig 3:**
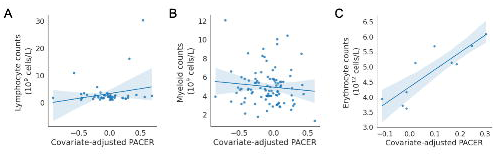
Scatterplot of blood counts versus covariate-adjusted passenger associated clonal expansion rate (PACER): (a) lymphocyte count (10^9^ cells/L) for patients with lymphoid mCAs, (b) myeloid count (10^9^ cells/L) for patients with myeloid mCAs, and (c) erythrocyte count (10^12^ cells/L) for patients with copy-neutral loss of heterozygosity or loss of the p arm of chromosome 9.

Since mutations in *JAK2*, such as *JAK2V617F*, on chromosome 9p are known to cause polycythemia vera, we performed a multiple regression of age at time of blood draw, sex, and PACER to predict erythrocyte count for individuals with CN-LOH or loss of chromosome 9p. This model explained 91.6% of the variance in erythrocyte count, and PACER had a significant positive association with erythrocyte count (β = 0.0119, 95% CI [0.006, 0.018], p = 0.018) (**Fig 3c**).

### Germline genetic determinants of mCA fitness

The high variability in clonal expansion rate across a wide range of individuals with the same mCA demonstrates that other factors, including germline mutations and environmental exposures likely affect mCA clonal expansion rate. To identify germline variants associated with the per-individual fitness estimated by PACER, we performed a GWAS of total passenger mutations in 6,381 individuals with 1 mCA (including mCAs of chromosome X) and without known CH driver SNVs (**Fig 4a**). We controlled for age, sex, ancestry, study cohort, and clonal fraction.

**Fig 4:**
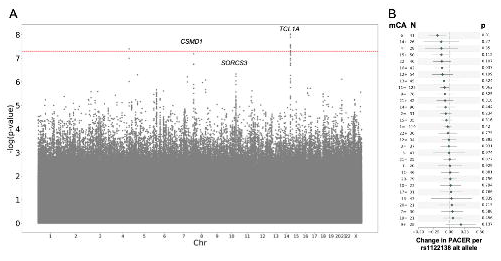
(a) Manhattan plot from the genome-wide association study (GWAS) for passenger- approximated clonal expansion rate (PACER) for single nucleotide polymorphisms (SNPs) with a minor allele frequency > 1%. A linear mixed model with kinship adjustment was used to regress total passenger mutations, with age, sex, clonal fraction, study, and the first 10 ancestral principal components included in the model as covariates. The dashed red line represents a p- value cutoff of 5×10^−8^. Nearest gene is labeled. SNPs within TCL1A had a p-value of 3.1×10^−8^, those within CSMD1 had a p-value of 1.4×10^−7^, and those within SORCS3 had a p-value of 4.6×10^−7^. (b) Forest plot of coefficients +/- 1.96 * SE and p-value for rs122138 alternate allele count in a multiple regression model of age at blood draw, sex, and clonal fraction to predict PACER, with number of individuals with that mCA labeled as “N”.

The GWAS identified a single nucleotide polymorphism (SNP) on chromosome 14, rs1122138, with genome-wide significance (p-value = 3.1×10^−8^). SNP rs1122138 is in an intronic region of *TCL1A*, and the alternate A allele is common, occurring in 21% of the haplotypes sequenced in TOPMed. However, rs1122138 and another SNP in the core promoter of *TCL1A*, rs2887399, which has been reported to modulate stem cell expansion for SNV CH,^20^ are in high linkage disequilibrium (R^2^ = 0.826) in the 1,000 Genome Project. The risk alleles, rs1122138(A) and rs2887399(T), were correlated and our conditional analysis of our GWAS summary statistics demonstrated that rs1122138 did not remain significant after adjusting for the effect of rs2887399. This *TCL1A* locus was previously associated with cross sectional prevalence of mCAs, suggesting the underlying mechanism for this locus is related to clonal expansion.^21,22^

To identify possible mCA-specific effects of rs1122138 on PACER, we performed a multiple regression of age, sex, clonal fraction, and rs1122138 alternate allele count predicting PACER for individuals with each mCA type. The coefficient for the rs1122138 alternate allele count varied by mCA type but did not significantly associate with PACER for any mCA type after multiple hypothesis correction (**Fig 4b)**.

We then sought to use mCA PACER as a tool to identify whether clonal expansion represented the underlying mechanism for any other loci previously identified associated with mCA prevalence. We interrogated loci identified in our previously reported GWAS of expanded mCA clone size; expanded clones were defined as clonal fraction > 10% of blood. In addition to *TCL1A*, we identified that SNPs overlapping *NRIP1* and *TERT* had a p-value less than 1×10^−3^ in our mCA PACER analysis (**Fig 5a**). Among the reported hits in the clonal expansion GWAS, both rs2887399 in *TCL1A* and rs2853677 in *TERT* were significantly negatively associated with PACER in our GWAS (**Fig 5b**).

**Fig 5:**
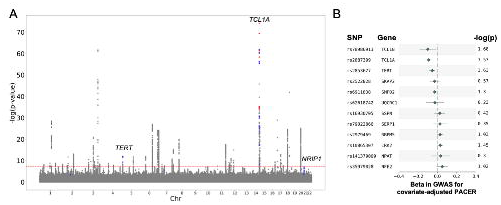
(a) Manhattan plot from the GWAS in Zekavat et al for expanded clonal size (defined as clonal fraction > 10%). Red points represent SNPs in the expanded clonal size GWAS with a p-value < 10^-6^ in the GWAS for passenger-approximated clonal expansion rate, and blue points represent SNPs in the expanded clonal size GWAS with a p-value < 10^-3^ and > 10^-6^ in the GWAS for passenger-approximated clonal expansion rate. These included SNPs in the gene TCL1A (chr14), TERT and NRIP1 (chr21). (b) Forest plot of the most significant SNPs in the GWAS for expanded clonal size demonstrating their coefficients and -log(p-value) in the GWAS of passenger-approximated clonal expansion rate. Genes annotated based on OpenTargets variant annotations. SNPs with beta < 0.025 not shown.

To identify rare germline variants associated with mCAs, we performed combined burden and variance-component tests implemented in REGENIE. We restricted our analysis to rare variants in coding regions of autosomal chromosomes (see **Methods**). We tested for associations with 17,668 genes, using five annotation masks, four of which predicted loss of function. No genes were significantly associated with mCAs (**Extended Data Fig 3**).

## Discussion

CH, which can be caused by SNVs or mCAs, is a common age-related condition that increases risk for hematologic malignancy, cardiovascular disease, liver disease, and all-cause mortality.^3,10,11,23–25^ The factors underlying clonal expansion in CH caused by mCAs have not been studied and are poorly understood. Here, we estimated the rate of clonal expansion in individuals with gain, loss, and CN-LOH mCAs in TOPMed. The PACER method has been previously validated in SNVs that cause clonal hematopoiesis of indeterminate potential (CHIP).^20^ Here we extend the approach to a distinct form of somatic mosaicism highlighting the generalizability of the approach and permitting several observations.

First, not only did the mCA clonal expansion rate vary significantly across chromosomes but also among individuals with the same driver mCA, highlighting that the driver mCA mutation was an incomplete determinant of clonal fitness. When we aggregate our fitness estimates by mCA location and type, the fitness estimated from passenger mutations correlated with an orthogonal approach by Watson and Blundell in the UK Biobank. Watson and Blundell use an evolutionary model of HSC dynamics to estimate the fitness of mCAs based on the clonal fraction distribution for individuals with the mCA.^18^ However, our method uniquely estimates the expansion rate for a clone within an individual, enabling single-variant analysis to find germline risk factors of clonal expansion and associations of fitness with clinical phenotypes, such as peripheral blood counts. The correlation observed between passenger-estimated fitness and CF-derived fitness suggests that PACER can provide per-individual fitness estimates comparable to fitness estimated with population-genetics methods.

Second, the fitness of mCAs estimated with PACER was lower than that of somatic SNV mutations in leukemia driver genes that cause CHIP. While CHIP mutations affect a single gene, mCAs typically span large genetic regions affecting the gene dose of dozens or even hundreds of genes. It is likely that these gene sets confer a mixture of both selective advantages balanced by deleterious consequences on the clonal outgrowth leading to overall decreased fitness compared to single gene mutations.

Third, we identified specific germline genetic determinants that contribute to an individual’s mCA expansion rate. We performed the first ever GWAS for germline modulators of mCA clone expansion rate. The GWAS, which identified *TCL1A* locus at genome-wide significance, suggests aberrant activation of TCL1A may also promote clonal expansion for mCAs as it does for CHIP. TCL1A is known to be part of the PI3K-Akt-mTOR signaling pathway. Previous work has proposed that the acquisition of a driver mutation can increase the accessibility of the pro-proliferative TCL1A gene, which promotes clonal expansion.^26^ This observation is convergent with prior observations that somatic rearrangements in TCL1A are implicated in lymphoid malignancies (specifically T-prolymphocytic leukemia).^20,27,28^ The results of this study provide further support for the role of TCL1A in clonal expansion.

We also leveraged our mCA expansion GWAS to identify NRIP1 and TERT as modulating mCA prevalence by affecting clonal expansion rate. NRIP1 is a regulator of oncogenic signaling pathways in chronic lymphocytic leukemia and a therapeutic target to sensitize acute myeloid leukemia to all-trans retinoic acid.^29,30^ Alternative SNPs in the *NRIP1* gene have been previously associated with increased WBC and monocyte count.^31^ TERT is a ribonucleoprotein polymerase that maintains telomere ends by addition of the TTAGGG repeat; its dysregulation in somatic cells is associated with oncogenesis. Rare variant association tests did not identify additional modulators of mCA clone expansion rate. Thus, our genetic analyses identify clonal expansion as a putative mechanism for selected loci associated with expanded mCA clone size.

Fourth, we find that mCA clonal expansion rate had phenotypic consequences. Previous work has identified an association among mCAs, white blood cell counts and infection rate, indicating that clonal expansion may lead to decreased ability to fight infection.^21^ We observe that faster mCA clonal expansion rate was associated with increased measured lymphocyte counts among individuals with mCAs. As the presence of an elevated lymphocyte count in combination with an mCA represents chronic lymphocytic leukemia, our observation suggests that mCA expansion rate in clones with lymphoid driver mutations may have utility in prognosticating risk of chronic lymphocytic leukemia progression. We also observe that higher clonal expansion rate is associated with increased erythrocyte counts in individuals with chromosome 9p mCAs affecting *JAK2*, a known gene implicated in polycythemia vera.

While our study provides novel insights into the fitness and germline modulators of mCAs, it has several limitations. First, TOPMed has a limited sample size of individuals with mCAs (6,381 individuals), so most mCAs were not present in more than 25 individuals and thereby were underpowered. Second, we were only able to study individuals with a single mCA due to limitations of the PACER method. Third, our results suggest that for some outlier mCAs, passenger mutations may be overestimated on the chromosome with the mCA due to structural rearrangements; however, this limitation does not seem to meaningfully affect our results.

Fourth, we do not currently have serial samples of measured mCA clonal fraction, so we are currently unable to validate our estimates of expansion rate. However, PACER estimates of clonal expansion have been successfully validated in individuals with SNV driver mutations.^20^

In summary, leveraging the per-individual mCA fitness estimations from PACER, we compared aggregate fitness of different mCA types and locations, identified germline determinants of mCA expansion rate and phenotypic consequences. These findings highlight potential treatment targets for mCA expansion rate and provide an approach to identify individuals at the highest risk of mCA-driven disease progression.

## Methods

### Study samples

For this study, we leveraged the NHLBI Trans-Omics for Precision Medicine (TOPMed) dataset, which has whole-genome sequencing on 127,946 samples from 51 studies with informed consent. The characteristics of this sample have been previously described.^20^ The study design was approved by the Vanderbilt Institutional Review Board (IRB#210270).

### Identification of mosaic chromosomal alterations with MoChA

Using WGS data from TOPMed, we identified 6,381 individuals with mosaic chromosomal alterations using MoChA.^32^ MoChA is a method that identifies mCAs using three hidden Markov models to find mCA-induced deviations in allelic balance at heterozygous sites, as described previously.^5,22^ An mCA was defined as a gain, loss, or copy-neutral loss of heterozygosity in a specific chromosome and p or q arm. Code is available at https://github.com/freeseek/mocha. We excluded 160 samples with phased B-allele frequency (BAF) auto-correlation >0.05, indicative of contamination or other potential sources of poor DNA quality, and 67 samples with phenotype-genotype sex discordance. We removed likely germline copy number polymorphisms (*lod_baf_phase* <20 for autosomal variants and *lod_baf_phase* <5 for sex chromosome variants), constitutional or inborn duplications (mCAs 2-10 Mb with relative coverage >2.25, and mCAs 50-250 Mb with relative coverage >2.5) and deletions (filtering out mCAs with relative coverage <0.5).

### Whole genome processing, variant calling, and exclusion of individuals with CHIP and multiple mCAs

Using previously described methods,^20^ we were able to call somatic singletons by identifying somatic SNVs that appeared in individuals with mCAs. Variants with a depth below 25 or above 100 were excluded, along with variants with a variant allele frequency exceeding 35% to exclude germline mutations. Individuals with mutations in genes associated with clonal hematopoiesis of indeterminate potential were excluded (e.g., *DNMT3A, ASXL1, TET2, JAK2*), as were individuals with multiple mCAs. The total number of “passenger mutations” – C>T or T>C base pair substitutions – was calculated for each patient with a single mCA.

### Passenger-approximated clonal expansion rate (PACER)

As was done in Weinstock et al,^20^ passenger mutations were defined as C>T and T>C base-pair substitutions that do not occur in a CH-associated gene. These mutations are “clock-like” and are associated with aging, whereas other mutations may be affected by disease or environmental factors. We can assume that all passenger mutations present at a WGS-detectable fraction occurred before the driver mCA because these ancestral passenger mutations will have increased abundance compared to sub-clonal passenger mutations.

### Covariate adjustment and normalization of total passenger mutation counts

After the number of total passenger mutations was calculated, we fit a negative binomial regression model of age, sex, and clonal fraction to predict total passenger mutations using scikit-learn in Python. We then performed a Yeo-Johnson inverse-normal transformation on the residuals using the SciPy package in Python 2.7.17. We then used these values as covariate-adjusted passenger-approximated clonal expansion rate to compare mCA fitness. We also calculated a covariate-adjusted PACER from passenger mutations excluding the chromosome of the mCA as described above. We performed a linear regression to compare covariate-adjusted PACER from passenger mutations including and excluding the mCA chromosome.

### Association between clonal expansion rate and blood counts

Peripheral counts for leukocytes, lymphocytes, neutrophils, basophils, eosinophils, monocytes, platelets, and erythrocytes were obtained from the TOPMed dataset for each individual with an mCA, along with the age of the person at the time of blood draw. The blood draw for sequencing and blood counts was within 2 years for all patients, and for 67% it was the same blood draw.

Myeloid cell counts were determined by summing counts of neutrophils, basophils, eosinophils, and monocytes. We employed previously defined curated sets of mCAs known to be associated with lineage-specific hematologic malignancies.^14,15^ Lymphoid mCAs included gain of chromosome 12, loss of the q arms of chromosomes 10 and 13, and CN-LOH of the q arms of chromosomes 8, 9 and 13. Myeloid mCAs included loss of the q arms of chromosomes 20 and 5, gain of chromosome 8, and CN-LOH of the q arms of chromosomes 9, 14, and 22. mCAs associated with polycythemia vera were defined as CN-LOH or loss of the p arm of chromosome 9. We used a two-tailed t-test to assess for differences in lymphocyte counts between lymphoid mCAs and all other mCAs and myeloid cell counts between myeloid mCAs and all other mCAs. We then used ordinary least squares regression to perform a multiple regression of age at time of blood draw, sex, and PACER to predict lymphocyte count among individuals with lymphoid mCAs, myeloid cell counts for individuals with myeloid mCAs, and erythrocyte counts for individuals with mCAs associated with polycythemia vera.

### Single variant association

Single variant association for each variant with minor allele frequency greater than 1% in individuals with a single mCA was performed with SAIGE.^33,34^ Analysis was performed using the TOPMed Encore analysis server (https://encore.sph.umich.edu). Covariates in the model were age at blood draw, sex, clonal fraction, TOPMed study, and the first ten genetic ancestry principal components. We applied an inverse normal transformation to the passenger counts. We declared variants from this analysis as significant if their p-value was less than 5 x 10^-8^.

### Linkage disequilibrium and conditional analysis for rs1122138

To determine whether rs1122138 is a distinct signal from rs2887399, a previously reported SNP associated with clonal expansion of SNV CH,^20^ we used the LDpair tool on LDLink (https://ldlink.nci.nih.gov). The R^2^ was used to assess for linkage disequilibrium. Then, we used PLINK to perform a conditional analysis to assess whether the association signal at rs1122138 remains significant after adjusting for the effect of rs2887399.^35^

### Rare single variant association

For the rare variant analysis, the omnibus test SKATO was selected because it combines variance component tests and burden tests. This analysis was implemented using a Regenie v3.2 pipeline, using the docker image released by the software creators.^36^ The covariates for steps 1 and 2 were age at blood draw, inferred sex, and the first ten principal components of genetic ancestry. Step 1 was restricted to a random selection of 500,000 extremely common variants.

Step 2 variants were rare (MAF < 0.01), in coding regions with mask annotations: nonsynonymous, stop-gain, stoploss, splicing, and exonic. The Bonferroni corrected significance threshold was 0.05/102140 ≈ 4.09×10^−7^.

## Supporting information

Supplementary Materials and Acknowledgements

## Acknowledgments

WGS for the Trans-Omics in Precision Medicine (TOPMed) program was supported by the National Heart, Lung, and Blood Institute (NHLBI). See the Supplementary Materials for study omics support information. Centralized read mapping and genotype calling, along with variant quality metrics and filtering, were provided by the TOPMed Informatics Research Center (3R01HL-117626-02S1; contract HHSN268201800002I). Phenotype harmonization, data management, sample identity quality control, and general study coordination were provided by the TOPMed Data Coordinating Center (R01HL-120393; U01HL-120393; contract HHSN268201800001I). We thank the studies and participants who provided biological samples and data for TOPMed. The full study-specific acknowledgments are included in the Supplementary Materials. The views expressed in this manuscript are those of the authors and do not necessarily represent the views of the National Heart, Lung, and Blood Institute; the National Institutes of Health; or the U.S. Department of Health and Human Services. We wish to acknowledge the contributions of the consortium working on the development of the NHLBI BioData Catalyst ecosystem.

## Data availability

Individual whole-genome sequence data for TOPMed whole genomes, individual-level harmonized phenotypes and the CHIP variant call sets used in this analysis are available through restricted access via the dbGaP TOPMed Exchange Area available to TOPMed investigators.

Controlled-access release to the general scientific community via dbGaP is ongoing. dbGaP accession numbers are available in Supplementary Tables 19 and 20 in Weinstock et al, 2023.^20^ GWAS summary statistics for PACER amongst individuals with mCAs are available in the following GitHub repository here: https://github.com/bicklab/pacer-mca-fitness/blob/main/Data/gwas_results_pacerint_nochip_df.assoc.

## Code availability

The code to call mosaic chromosomal alterations with MoChA is available here: https://github.com/freeseek/mocha. Passenger count variant calling pipeline is available here: https://github.com/weinstockj/passenger_count_variant_calling. Code for the analyses of this paper can be found here: https://github.com/bicklab/pacer-mca-fitness.

---

**Extended Data Fig 1:**
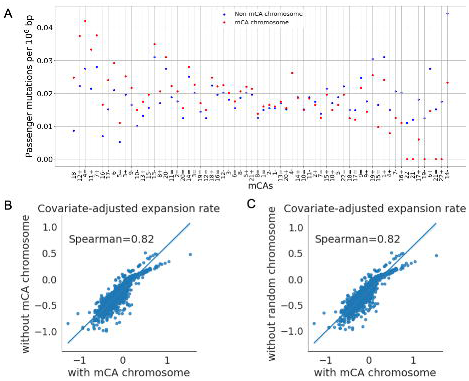
**(a)** Scatterplot of median passenger mutation density per 106 base pairs (bp) by mCA for chromosomes without the mCA (blue) and with the mCA (red). The x-axis is ordered from greatest to least difference between the passenger mutation density on mCAs and non-mCAs. “+” represents gain of chromosome, “-“ represents loss of chromosome, and “=” represents CN-LOH. **(b-c)** Correlation between the difference in passenger mutation density between mCA and non-mCA chromosomes and covariate-adjusted passenger-approximated clonal expansion rate (PACER). The R^2^ value of PACER and the difference in passenger mutations per 10^6^ bp is 0.16.

**Extended Data Fig 2:**
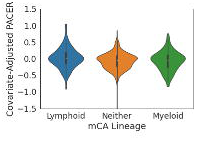
Covariate-adjusted PACER score in individuals with a lymphoid-associated mCA compared to myeloid associated mCAs and mCAs with no particular association. The PACER clonal expansion rate is calculated after covariate adjustment (age, sex, clonal fraction) and inverse normalization of the total passenger mutations for a given individual.

**Extended Data Fig 3:**
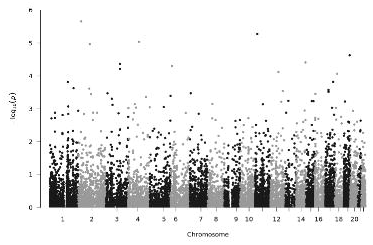
Manhattan plot from the rare variant analysis of PACER, consisting of combined burden and variance-component tests implemented in REGENIE. We restricted our analysis to rare variants in coding regions of autosomal chromosomes. We tested for associations with 17,668 genes, using five annotation masks. Further adjusting for 5 annotation masks per gene results in a strict Bonferroni adjusted significance threshold of p < 5.7x 10^-7^. No rare variants were statistically significant.

